# Balanced steady state free precession enables high-resolution dynamic 3D Deuterium Metabolic Imaging of the human brain at 7T

**DOI:** 10.1101/2025.02.06.25321580

**Authors:** Sabina Frese, Bernhard Strasser, Lukas Hingerl, Elton Montrazi, Lucio Frydman, Stanislav Motyka, Viola Bader, Anna Duguid, Aaron Osburg, Martin Krssak, Rupert Lanzenberger, Thomas Scherer, Wolfgang Bogner, Fabian Niess

**Affiliations:** High Field MR Center, Department of Biomedical Imaging and Image-Guided Therapy, Medical University of Vienna, Lazarettgasse 14, Vienna A-1090, Austria; Department of Chemical and Biological Physics, Weizmann Institute of Science, Rehovot, Israel; Christian Doppler Laboratory for MR Imaging Biomarkers (BIOMAK), Austria; Department of Medicine III, Division of Endocrinology and Metabolism, Medical University of Vienna, Austria; Department of Psychiatry and Psychotherapy, Comprehensive Center for Clinical Neurosciences and Mental Health (C3NMH), Medical University of Vienna, Austria

**Keywords:** Deuterium Metabolic Imaging, whole-brain metabolic mapping, deuterium-labeled glucose, 3D magnetic resonance spectroscopic imaging, balanced steady state free precession, downstream neurotransmitter synthesis, continuous glucose monitoring

## Abstract

**Objectives:** Deuterium (^2^H) Metabolic Imaging (DMI) is an emerging magnetic resonance technique to non-invasively map human brain glucose (Glc) uptake and downstream metabolism following oral or intravenous administration of ^2^H-labeled Glc. The achievable spatial resolution is limited due to inherently low sensitivity of DMI. This hinders potential clinical translation. The purpose of this study was to improve the signal-to-noise ratio (SNR) of 3D DMI via a balanced steady state free precession (bSSFP) acquisition scheme combined with fast non-Cartesian spatial-spectral sampling to enable high resolution dynamic imaging of neural Glc uptake and glutamate+glutamine (Glx) synthesis of the human brain at 7T.

**Materials and Methods:** Six healthy volunteers (2f/4m) were scanned after oral administration of 0.8 g/kg [6,6’]-^2^H-Glc using a novel density-weighted bSSFP acquisition scheme combined with fast 3D concentric ring trajectory (CRT) k-space sampling at 7T. Time-resolved whole brain DMI datasets were acquired for approximately 80 min (7 min per dataset) after oral ^2^H-labeled Glc administration with 0.75ml and 0.36ml isotropic spatial resolution and results were compared to conventional spoiled Free Induction Decay (FID) ^2^H-MRSI with CRT readout at matched nominal spatial resolution.

Dynamic DMI measurements of the brain were accompanied by simultaneous systemic Glc measurements of the interstitial tissue using a continuous Glc monitoring (CGM) sensor (on the upper arm). The correlation between brain and interstitial Glc levels was analyzed using linear mixed models.

**Results:** The bSSFP-CRT approach achieved SNRs that were up to 3-fold higher than conventional spoiled FID-CRT ^2^H-MRSI. This enabled a 2-fold higher spatial resolution. Seventy minutes after oral tracer uptake comparable ^2^H-Glc, ^2^H-Glx and ^2^H-water concentrations were detected using both acquisition schemes at both, regular and high spatial resolutions (0.75ml and 0.36 ml isotropic). The mean Areas Under the Curve (AUC) for interstitial fluid Glc measurements obtained using a continuous Glc monitoring (CGM) sensor was 509±65 mM·min. This is 3.4 times higher than the mean AUC of brain Glc measurements of 149±43 mM·min obtained via DMI. The linear mixed models fitted to assess the relationship between CGM measures and brain ^2^H-Glc yielded statistically significant slope estimates in both GM (β_1_ = 0.47, *p* = 0.01) and WM (β_1_ = 0.36, *p* = 0.03).

**Conclusion:** In this study we successfully implemented a balanced steady state free precession (bSSFP) acquisition scheme for dynamic whole-brain human DMI at 7T. A 3-fold SNR increase compared to conventional spoiled acquisition allowed us to double the spatial resolution achieved using conventional FID-CRT DMI. Systemic continuous glucose measurements, combined with dynamic DMI, demonstrate significant potential for clinical applications. This could help to improve our understanding of brain glucose metabolism by linking it to time-resolved peripheral glucose levels. Importantly, these measurements are conducted in a minimally invasive and physiological manner.

## Introduction

The mammalian brain relies mainly on glucose (Glc) to fuel neuronal activity, neurotransmitter release and brain cell communication (1, 2). Impaired brain Glc uptake and metabolism are hallmarks of many neurological pathologies including neurodegenerative disorders and brain tumors, as well as of metabolic diseases such as diabetes (3–6).

The gold standard imaging technique for tissue-specific Glc uptake is Fluorodeoxyglucose positron emission tomography ([^18^F]-FDG-PET), where the radioactive decay of the [^18^F]- isotope allows localization of Glc uptake. Due to glucose trapping, the tracer accumulates in the tissue over time, indirectly representing metabolic activity. However, FDG-PET, can neither provide direct information on downstream metabolism nor differentiate between oxidative and glycolytic pathways (7, 8).

Deuterium (^2^H) Metabolic Imaging (DMI) is an emerging magnetic resonance (MR) technique that enables the non-invasive imaging of Glc uptake, as well as downstream metabolism into neurotransmitters such as glutamate and glutamine (Glx) or lactate (Lac). DMI uses non-radioactive, minimally toxic, and stable ^2^H-labeled glucose solutions (9–12). Further advantages of DMI compared to more conventional ^1^H-MR spectroscopic imaging (MRSI) approaches are spectral sparsity – i.e., the targeting of a low number of *a priori* known, spectrally-resolved metabolites – and a low natural abundance (0.0156%) of deuterium for the *in vivo* water and lipid resonances. This renders water and lipid suppression unnecessary and simplifies internal water referencing. Still, clinical translation of DMI is challenged by an inherently low sensitivity and prolonged acquisition times. Both limit the achievable spatial resolution, which is currently well below the current gold standard FDG-PET.

Recently, fast spatial-spectral k-space sampling schemes using non-Cartesian trajectories have been implemented for DMI applications in the human brain (13) and liver (14). These enabled an increased spatial resolution compared to conventional phase-encoded sampling, without prolonging scan duration. The ensuing smaller voxel sizes reduce the signal-to-noise ratio (SNR), but this was counterbalanced by using low-rank denoising algorithms during post processing (13, 14). In the brain, higher spatial resolution enabled improved metabolic differentiation of gray and white matter (GM/WM) (13). However, further increases in spatial resolution to better resolve smaller local metabolic alterations or detect lower concentrations, is still a challenge. Particularly valuable would be to quantify the early metabolic turnover rates of Glc directly after tracer uptake, and early dynamics that are hard to quantify when the *in vivo* concentrations of the labeled downstream metabolites are still relatively low.

To increase sensitivity and consequently the spatial resolution, balanced steady state free precession (bSSFP) acquisition schemes can be employed (15, 16). Due to the quadrupolar nature of deuterium, relaxation times T_1_ and T_2_ of DMI’s metabolic targets are quite similar, making DMI a well-suited candidate for acquisitions using bSSFP sequences. Employing such acquisition schemes, recent preclinical DMI studies have shown up to 4-fold SNR increases at 15.2T (17–19).

The aim of this study was to develop a bSSFP sequence combined with a CRT readout, to boost SNR and thereby enable dynamic high-resolution whole-brain DMI in humans at 7T. We demonstrate the feasibility of bSSFP-CRT to map the healthy glucose metabolism with 2-fold increased spatial resolution, and evaluated the reproducibility to estimate dynamic concentrations of ^2^H-Glc and ^2^H-Glx compared to more conventional FID-CRT acquired during the same session.

## Materials and Methods

### Study participants

Six healthy volunteers (age: 27±2 years; Body Mass Index (BMI): 24±2 kg/m2, 4 male/2 female) participated in the study after written informed consent was obtained (Table 1). The study was approved by the local ethics committee of the Medical University of Vienna according to the guidelines of the Declaration of Helsinki. To assess baseline glucose levels before, during and after the DMI experiment all participants were asked to wear an MR-safe device for continuous glucose measurement (CGM) of the interstitial tissue on the upper arm (FreeStyle Libre®3, Abbott Laboratories), applied the day before. The vendor’s intended use does not officially include MR-approval, but the sensor was extensively tested for MR-safety in a recent study (20). For the current study, the local ethics committee approved the use of the device for investigational purposes.

**Table 1:**
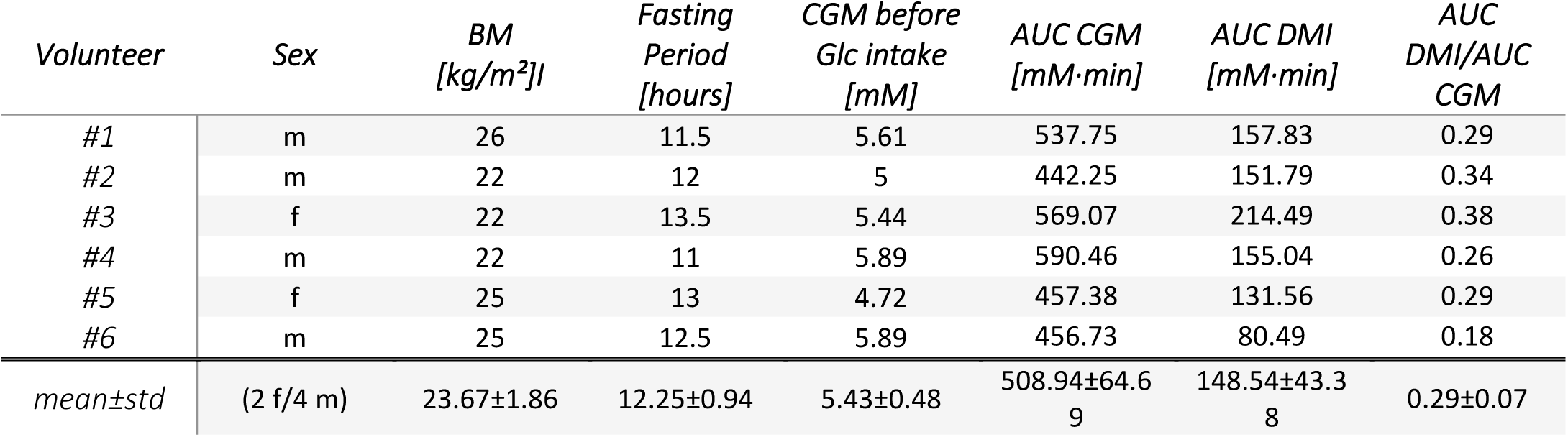
Summary of the 6 healthy volunteers included in this study, with respective sex, Body Mass Index (BMI), fasting period in hours, continuous glucose monitoring (CGM) values before ^2^H-labeled glucose (Glc) intake, as well as areas under the curve (AUC) determined with CGM and deuterium metabolic imaging (DMI) and their ratio, and mean ± standard deviation (std) over all subjects.

| *Volunteer* | *Sex* | *BM*<br>*[kg/m<sup>2</sup>]* | *Fasting*<br>*Period*<br>*[hours]* | *CGM before*<br>*Glc intake*<br>*[mM]* | *AUC CGM*<br>*[mM·min]* | *AUC DMI*<br>*[mM·min]* | *AUC*<br>*DMI/AUC*<br>*CGM* |
| --- | --- | --- | --- | --- | --- | --- | --- |
| #1 | m | 26 | 11.5 | 5.61 | 537.75 | 157.83 | 0.29 |
| #2 | m | 22 | 12 | 5 | 442.25 | 151.79 | 0.34 |
| #3 | f | 22 | 13.5 | 5.44 | 569.07 | 214.49 | 0.38 |
| #4 | m | 22 | 11 | 5.89 | 590.46 | 155.04 | 0.26 |
| #5 | f | 25 | 13 | 4.72 | 457.38 | 131.56 | 0.29 |
| #6 | m | 25 | 12.5 | 5.89 | 456.73 | 80.49 | 0.18 |
| *mean±std* | (2 f/4 m) | 23.67±1.86 | 12.25±0.94 | 5.43±0.48 | 508.94±64.6<br>9 | 148.54±43.3<br>8 | 0.29±0.07 |

After overnight fasting (12±1 hours), participants orally consumed deuterium-labeled (^2^H) Glc (0.8 g/kg body weight, [6,6’]-^2^H-Glc ≥99% purity, Cambridge Isotopes) dissolved in ∼200 ml water, immediately before they were moved inside the magnet bore.

### Deuterium metabolic imaging

Measurements were performed on a human whole-body 7T (Magnetom dot Plus) Siemens MR system using a ^2^H/^1^H dual-tuned quadrature birdcage head coil (Stark Contrasts MRI Coils Research, Germany). Scanner hardware or software modification was not required as the acquisition of ^2^H signals is supported by the vendor.

#### Sequence development

Based on a previously developed spoiled FID ^2^H-MRSI sequence (13), a bSSFP acquisition scheme was implemented and combined with a fast 3D Hamming-weighted concentric ring trajectory readout (CRT). bSSFP parameter estimation was performed as previously described (17) and adapted for measurements in the human brain at 7T to optimize for detection of ^2^H resonances of water (4.8 ppm), Glc (3.9 ppm), Glx (2.4 ppm) and Lac (1.3 ppm), yielding the following imaging parameters: T_R_ = 23 ms, flip angle: 50° (phase cycling factor 2), block pulse duration: 1.0 ms, acquisition delay: 2.0 ms, echo spacing ΔT_E_ = 3.5 ms, central frequency: 3.2 ppm, receiver bandwidth 285 Hz, 5 sample points, N_rings_=107 (for density weighted averaging), 80 preparation scans to achieve a steady state magnetization (Figure 1a).

**Figure 1:**
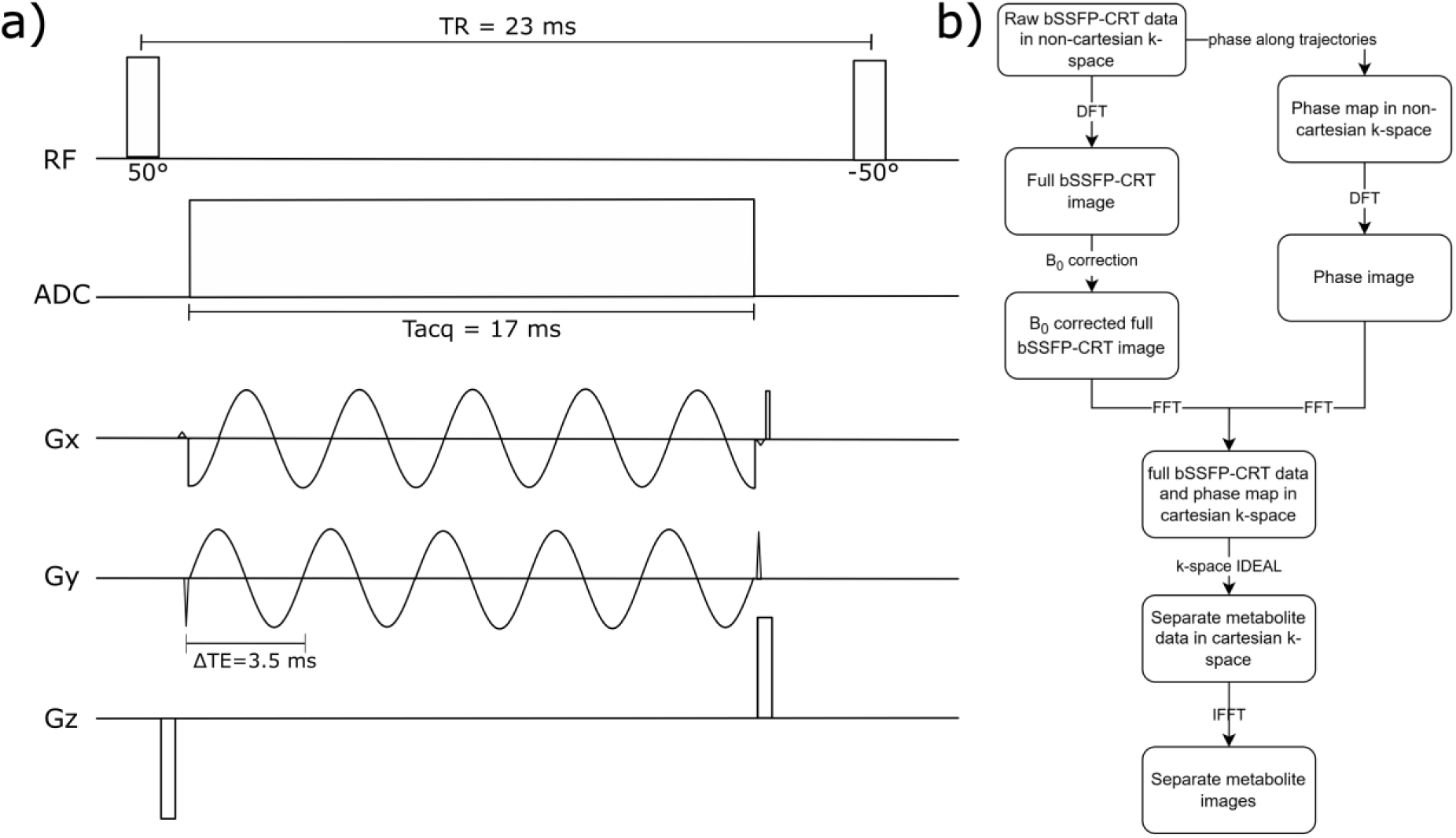
a) Pulse scheme of the new balanced Steady State Free Precession sequence with concentric ring trajectory readout (bSSFP-CRT) for DMI with the optimized parameters. To avoid banding artifacts the phase of the flip angle was alternated with each excitation by 180° yielding a flip angle of ±50°. b) Reconstruction workflow of the bSSFP-CRT DMI sequence: Raw non-cartesian data is read into MATLAB 2017. Before reconstruction using Discrete Fourier Transformations (DFT), phase maps were created along the concentric ring trajectories in non-cartesian k-space. After DFT, the data was corrected for B_0_ inhomogeneities with a B_0_ map acquired at the end of the DMI experiment. The B_0_ corrected image and the phase image are thereafter transformed to cartesian k-space using Fast Fourier Transformations (FFT), where metabolite separation is performed by accounting for phase evolution between the echoes as well as the points along the non-cartesian k-space trajectories. The separated metabolites are thereafter transformed to image space using an Inverse Fast Fourier Transformation (IFFT).

#### Phantom measurements

The SNR was assessed in a phantom containing three separate containers filled with 500 ml sodium chloride (0.9%), 50 ml aqueous ^2^H-glucose solution (0.3% [6,6’]-^2^H-Glc), and 1000 ml sodium lactate (60% INCI) solution giving rise to natural abundance levels of ^2^H-water, ^2^H-Glc and ^2^H-Lac, respectively. Data were acquired using the bSFFP-CRT sequence and compared to conventional spoiled FID-CRT acquisitions at matched spatial resolutions (FOV: 200×200×192 mm^3^, matrix: 22×22×21, 0.75 ml isotropic) and acquisition durations (7 min each). Data acquisition was repeated five times for both acquisition schemes and SNR was calculated as mean over the standard deviation for each separated metabolite map, i.e., water, glucose and lactate, across all repetitions in the respective containers of interest. Sequence parameters of the FID-CRT sequence were as follows: T_R_=290 ms, flip angle: 86°, block pulse duration: 1.0 ms, acquisition delay: 2.0 ms, central frequency: 4.8 ppm, receiver bandwidth: 380 Hz, 96 spectral points, N_rings_=43.

#### Dynamic brain DMI

Dynamic DMI measurements were performed following initial preparation scans, i.e., auto-align localizer images and unlocalized pulse-acquire B_1_ estimation (T_R_=1.5s, T_E_=0.35ms, 20 steps, U_Ref_=20-440 V). In all participants, ten whole-brain DMI datasets were measured consecutively over ∼70 min (∼7 min per 3D dataset), using both FID-CRT (first and last) and bSSFP-CRT (second to ninth) acquisition schemes.

Three participants were scanned at matched nominal spatial resolution (FOV: 200×200×192 mm^3^, matrix: 22×22×21, 0.75 ml isotropic) to validate the novel acquisition technique (Figure 2a), while for the other three participants, the spatial resolution of the bSSFP-CRT sequence was increased by 2-fold (FOV: 200×200×192 mm^3^, matrix: 28×28×27, 0.36 ml isotropic) (Figure 2a). One participant underwent an additional DMI scan at the end of the dynamic DMI protocol using the FID-CRT sequence with matched spatial resolution (0.36 ml isotropic).

**Figure 2:**
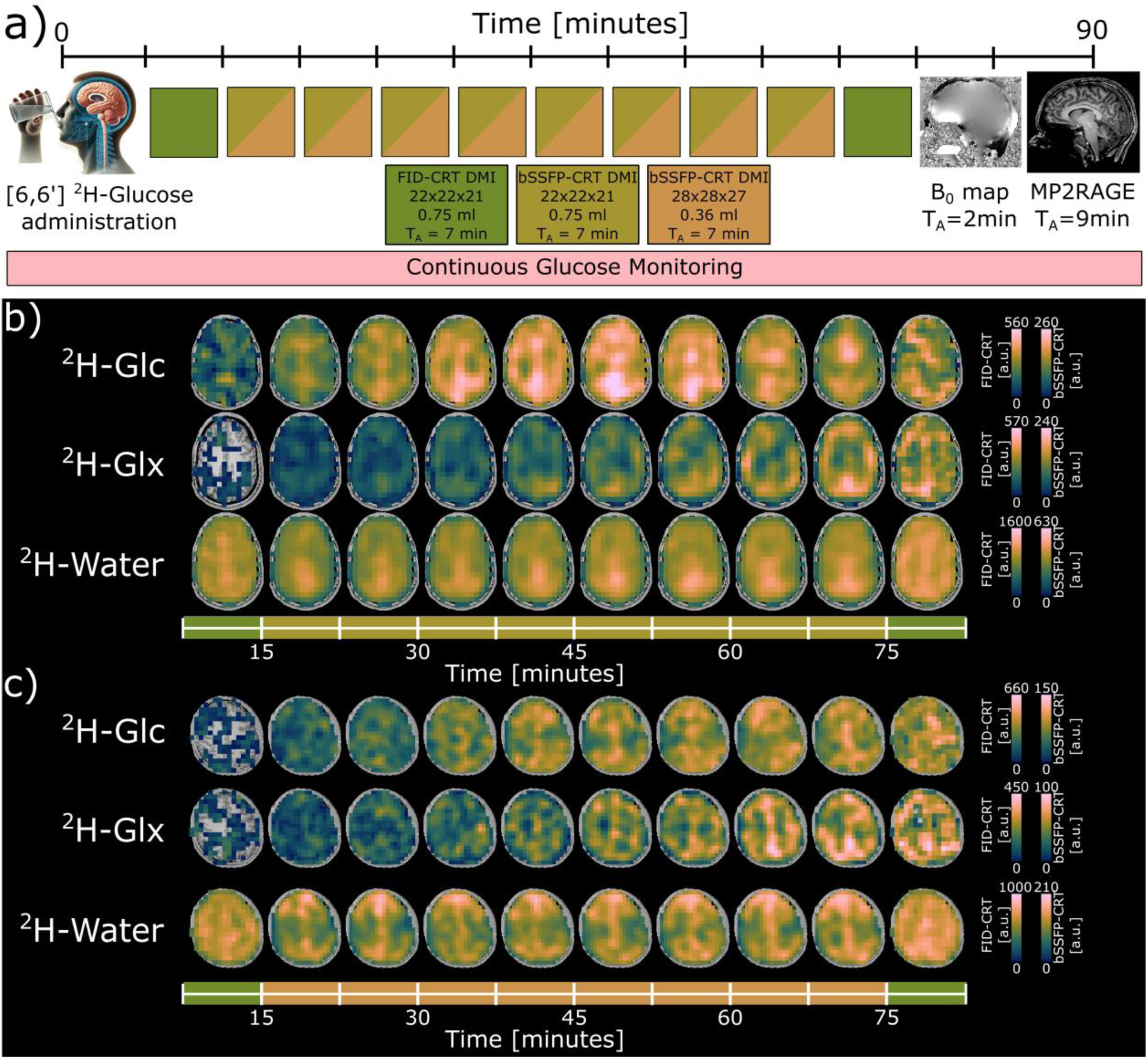
a) Schematic illustration of the experimental 7T scanning protocol for the healthy volunteers. The participant was placed into the magnet bore immediately after oral administration of [6,6’]-^2^H Glucose (Glc). Ten 3D whole-brain deuterium metabolic imaging (DMI) datasets were acquired over ∼70min after initial preparation scans using two acquisition schemes with matched acquisition duration (7 min), i.e., Free Induction Decay Concentric Ring Trajectory (FID-CRT) ^2^H-MRSI (0.75 ml isotropic resolution, dark green) and matched and increased resolution balanced Steady State Free Precession Concentric Ring Trajectory DMI (bSSFP-CRT, 0.75 ml (olive green) and 0.36 ml (orange) isotropic resolution) The DMI measurements were complemented by monitoring systemic Glc levels using a continuous glucose monitoring senor. Additionally, a high resolution B_0_-map and a T_1_-weighted anatomical image (MP2RAGE) were acquired. b) Time courses of axial ^2^H-Glc, ^2^H-glutamate+glutamine (Glx) and ^2^H-water maps in arbitrary units (a.u.) from one representative volunteer scanned with FID-CRT (dark green) and bSSFP-CRT (olive green) at matched spatial resolution (0.75ml isotropic). c) Time courses of axial ^2^H-Glc, ^2^H-glutamate+glutamine (Glx) and ^2^H-water maps in arbitrary units (a.u.) from one representative volunteer scanned with FID-CRT (0.75ml isotropic, dark green) and bSSFP-CRT (0.36ml isotropic, orange). Missing voxels do not contain a value (NaN: not a number).

Dynamic DMI measurements of all participants were followed by acquisitions of a high-resolution ^1^H-B_0_ map (FOV: 240×240×160 mm^3^, matrix: 128×128×80, T_R_=1410 ms, T_E1_=3 ms, T_E2_=6 ms, T_E3_=9 ms, T_E4_=12 ms, T_E5_=15 ms, T_A_=2:08 min (21)), and a high-resolution T_1_-weighted anatomical image (MP2RAGE; FOV: 165×220×220 mm^3^, matrix: 144×192×192, T_R_=3930 ms, T_I1_=850 ms, T_I2_=3400 ms, T_E_=3.28 ms, T_A_=9:29 min). The total duration of the DMI protocol was ∼90 min and was complemented by simultaneous CGM using a FreeStyle Libre®3 sensor.

Detailed information on sequence optimization and parameters is provided in Supplemental Digital Content (Figure 1 and Table 1 for minimum reporting standards (22)).

### B_0_ maps

^1^H-B_0_ maps were created with the ASPIRE algorithm (21), yielding voxel wise inhomogeneities in Hz. These maps were downsampled to the grid size of the DMI datasets (22×22×21/28×28×27) using the MINC toolbox (23). Thereafter, ^2^H-B_0_ maps were calculated according to Δ*f* = *γ* · Δ*B*_0_, where Δ*f* is the inhomogeneity in Hz, *γ* is the gyromagnetic ratio and· Δ*B*_0_ is the inhomogeneity in T.

### Data processing and metabolite quantification

Image reconstruction was performed offline using an in-house developed post-processing pipeline (MATLAB R2017) consisting of non-Cartesian three-dimensional discrete Fourier transformation applied after density correction to a 3D Hamming weighted k-space (24, 25). *In vivo* datasets from both DMI sequences were corrected for B_0_-inhomogeneities before spectral fitting or separating the ^2^H-resonances using the acquired ^2^H-B_0_ maps.

Spectral fitting of FID-CRT data was performed using LCModel with a simulated basis set (26, 27) including ^2^H-resonances of natural abundance water (4.8 ppm), Glc (3.9 ppm), Glx (2.4 ppm) and Lac+Lipid (1.3 ppm). As only healthy participants were included and, thus, low lactate levels were expected, this study focuses only on ^2^H-labeled water, Glc and Glx. For further analysis, quantitative results with a Cramer-Rao Lower Bound (CRLB) >50% were excluded.

Metabolic separation of the aforementioned resonances in the bSSFP-CRT datasets was performed using an adapted k-space IDEAL algorithm (28), to account for phase evolution during acquisition along the non-Cartesian concentric ring trajectories (Figure 1b).

### Segmentation

High resolution T_1_-weighted anatomical 3D images (MP2RAGE) were used for segmentation of gray and white matter (GM/WM) regions by calculating voxel-wise fraction maps of GM, WM and cerebrospinal fluid (CSF) using the FAST algorithm (29). Tissue-specific maps were downsampled to DMI grid size in the k-space to match the partial volume effects of the 3D Hamming filter. A threshold of 60% was used for regional averaging over GM and WM dominated regions in DMI datasets. Regional averaging over CSF was not performed due to its low relative content in downsampled tissue-specific maps.

### Concentration estimation

For all ten time points, ^2^H-Glc and ^2^H-Glx concentration estimates for GM and WM dominated regions were calculated using natural abundance water signals from the beginning of the dynamic DMI measurement as internal reference (30). Relaxation times and *in vivo* concentrations were assumed from literature (31, 32) (^2^H-water: T_1GM_/T_1WM_/T_1CSF_=358/328/510 ms, T_2GM_/T_2WM_/T_2CSF_=36/34/90 ms, ^2^H-Glc: T_1_/T_2_=80/34 ms, ^2^H-Glx: T_1_/T_2_=165/33 ms, pure ^2^H-water concentration: 17.2 mM), while taking into account different relaxation mechanisms in FID-CRT and bSSFP-CRT and the number of deuterium atoms per molecule (Supplemental Digital Content, Appendix 1). Voxel wise fractional water content of GM, WM and CSF was accounted for based on segmentation maps. ^2^H-Glx maps were corrected for an assumed 40% label loss in the tricarboxylic acid (TCA) cycle (33).

### Comparison between dynamic FID-CRT and bSSFP-CRT DMI

*In vivo* FID-CRT and bSSFP-CRT data were compared based on the metabolite concentrations of ^2^H-Glc and ^2^H-Glx measured at ∼70min (last bSSFP-CRT time point) and ∼77 min (last FID-CRT time point) after oral tracer uptake, respectively. To investigate regional differences in Glc metabolism arising from the acquisition scheme, the mean and standard deviation of ^2^H-Glc, ^2^H-Glx and ^2^H-water concentrations were calculated over GM and WM dominated regions for each volunteer and both acquisition schemes acquired at ∼70 min and ∼77 min after ^2^H-Glc administration.

Paired t-tests were conducted using R (version 4.1.2, 2021) to compare the concentrations of ^2^H-Glc, ^2^H-Glx and ^2^H-water between GM and WM regions, as well as between the last bSSFP-CRT and FID-CRT time points with a significance level of *p*<0.05 (34).

### Comparison between DMI and CGM

Two linear mixed-effects models (using R, version 4.1.2, 2021) were used to analyze the relationships between ^2^H-Glc concentrations measured in GM and WM dominated regions of the brain, and systemic Glc levels measured in the interstitial tissue using a CGM sensor (fixed effect). A random intercept and slope for each subject were fitted to account for inter-subject variability and differences between ^2^H-Glc levels in the brain and peripheral Glc levels. Significance level was set to *p*<0.05.

Additionally, area under the curve (AUC) values were calculated using trapezoid integration in R (version 4.1.2, 2021) for time courses of brain glucose levels detected via DMI and systemic glucose levels measured using the CGM sensor. The ratio of both glucose measurements was calculated for every participant.

## Data availability statement

Data generated by postprocessing (i.e., metabolic maps, LCModel basis sets, adapted k-space IDEAL algorithm, script files for data plotting) are available from the corresponding author on reasonable request for research purposes only.

## Results

### Deuterium metabolic imaging

#### Phantom measurements

The measured SNR using the bSSFP-CRT DMI sequence were ∼63 and ∼40 for ^2^H-water and ^2^H-Glc, respectively, whereas the SNR in the FID-CRT acquisition scheme were ∼23 and ∼13 for ^2^H-water and ^2^H-Glc, respectively. The SNR in ^2^H-Lac could not be reliably determined using both acquisition schemes. Consequently, the SNR gain of the bSSFP-CRT over the FID-CRT sequence was 2.74 and 3.07 for ^2^H-water and ^2^H-Glc, respectively.

#### Dynamic brain DMI

Initial preparation scans prior to the dynamic DMI protocol were completed within 8.83±1.47 min.

Time courses of axial metabolic maps of ^2^H-glucose (Glc), ^2^H-glutamate+glutamine (Glx) and ^2^H-water from one representative volunteer are shown in Figure 2b using both bSSFP-CRT and FID-CRT acquisition schemes with matched nominal spatial resolution (0.75 ml isotropic). bSSFP-CRT enabled a 2-fold increase in nominal spatial resolution (0.36 ml isotropic). Time courses of axial metabolic maps together with DMI maps acquired using FID-CRT (0.75 ml isotropic) during the same session from one representative volunteer are shown in Figure 2c. Individual maps from all participants are shown in Supplemental Digital Content (Figures 2-4) for ^2^H-Glc, ^2^H-Glx and ^2^H-water, respectively.

Direct comparison of 3D metabolic maps of ^2^H-glucose (Glc), ^2^H-glutamate+glutamine (Glx) and ^2^H-water acquired using bSSFP-CRT and FID-CRT acquisition schemes at matching resolutions from two representative volunteers are displayed in axial, sagittal and coronal views in Figure 3.

**Figure 3:**
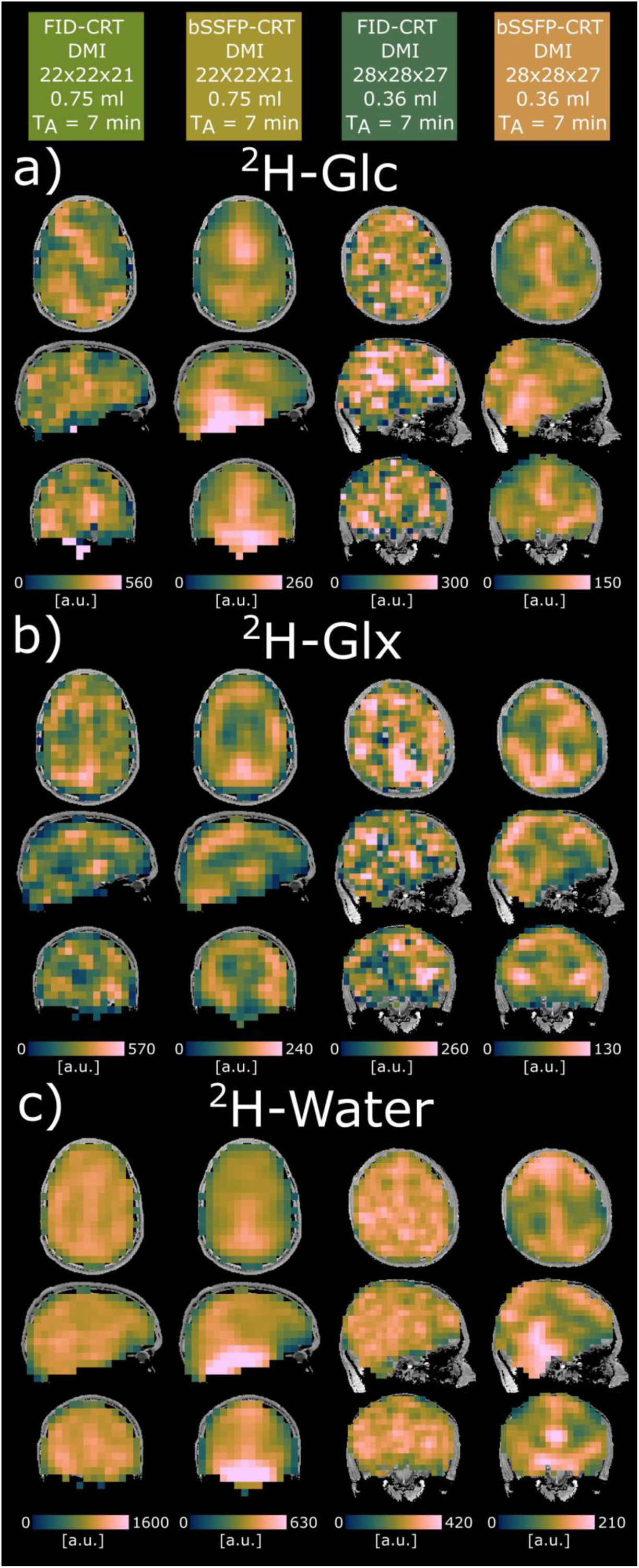
Representative metabolic maps of two subjects imaged with 0.75ml (left two columns) and 0.36ml (right two columns) isotropic spatial resolution with the two acquisition schemes, Free Induction Decay Concentric Ring Trajectory ^2^H-MRSI (FID-CRT) Deuterium Metabolic Imaging (DMI) and balanced steady state free precession concentric ring trajectory (bSSFP-CRT) DMI of ^2^H-Glucose (Glc, a), ^2^H-Glutamate+Glutamine (Glx, b) and ^2^H-water (c) in arbitrary units (a.u.). The images were acquired >1 hour after oral administration of ^2^H-Glc.

For all time points, mean concentration estimates of ^2^H-Glc, ^2^H-Glx and ^2^H-water over GM and WM dominated regions and across subjects with the same acquisition protocols are illustrated in Figure 4.

**Figure 4:**
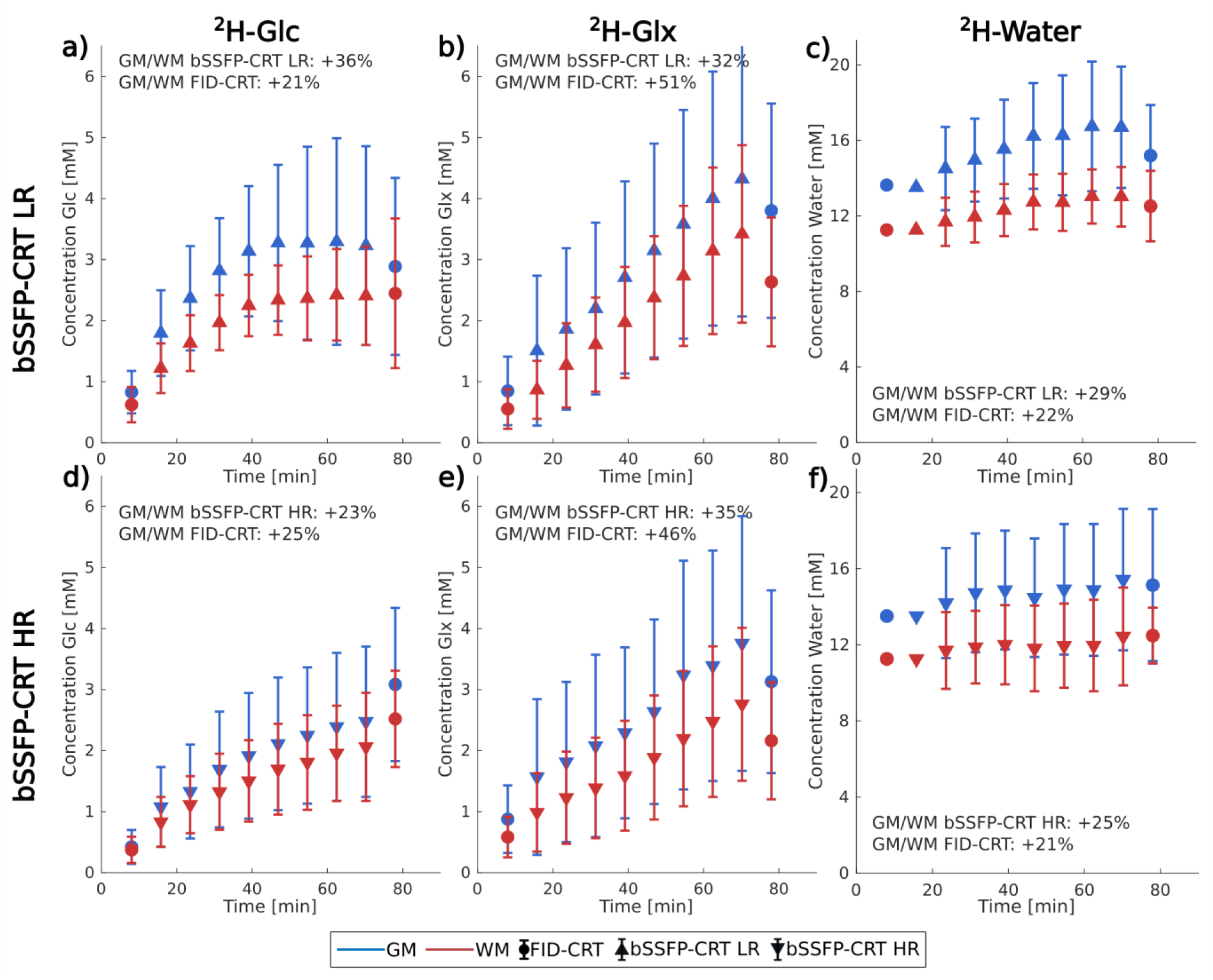
Time courses in mM of ^2^H-labeled glucose (Glc, a & d), glutamate+glutamine (Glx, b & e) and water (c & f), as regional means and the standard deviation over gray (GM, blue) and white matter (WM, red) over all subjects acquired with 0.75ml isotropic resolution in both Free Induction Decay Concentric Ring Trajectory (FID-CRT) and balanced Steady State Free Precession Concentric Ring Trajectory (bSSFP-CRT) DMI (upper row) and with 0.36ml isotropic resolution in bSSFP-CRT DMI (lower row). Over all participants, significant contrasts (GM/WM) were found in both acquisition schemes in water (bSSFP-CRT: +27%, *p*<0.05; FID-CRT: +22%, *p*<0.05), but not in ^2^H-Glc (bSSFP-CRT: +31%, *p*=0.12; FID-CRT: +22%, *p*=0.12) and ^2^H-Glx (bSSFP-CRT bSSFP-CRT: +34%, *p*=0.12; FID-CRT: +49%, *p*=0.05). No significant differences between the last time points of the acquisition schemes were observed.

### Comparison between dynamic FID-CRT and bSSFP-CRT DMI

Approximately 70 min after tracer intake averaged ^2^H-Glc concentration estimates derived from bSSFP-CRT DMI maps increased to 2.91±1.21 mM and 2.22±0.83 mM (mean±STD) in GM and WM dominated regions across all subjects, respectively. Similarly, averaged ^2^H-Glc concentration estimates derived from FID-CRT DMI maps in the same volunteers at ∼77 min after tracer administration were 2.96±1.22 mM and 2.42±1.04 mM in GM and WM, respectively. No significant differences were found between bSSFP-CRT and FID-CRT acquisition schemes in GM (*p*=0.14) and WM (*p*=0.22) regions. Higher ^2^H-Glc concentrations were observed in GM compared to WM dominated regions for both acquisition schemes (bSSFP-CRT: +31%, *p*=0.03; FID-CRT: +22%, *p*=0.03).

Averaged ^2^H-Glx concentrations measured with the bSSFP-CRT sequence ∼70 min after tracer intake increased to 4.05±0.93 mM and 3.05±0.71 mM in GM and WM, respectively. While averaged ^2^H-Glx concentration estimates derived from FID-CRT data ∼77 min after tracer uptake were similar in GM regions (3.42±0.82 mM, *p*=0.11), smaller concentrations were observed in WM regions (2.29±0.53 mM, *p*=0.04) compared to bSSFP-CRT data. Higher ^2^H-Glx concentrations were observed in GM compared to WM dominated regions for both acquisition schemes (bSSFP-CRT: +34%, *p*=0.04; FID-CRT: +49%, *p*=0.007).

^2^H-water concentration estimates derived from bSSFP-CRT maps were 16.13±3.09 mM and 12.71±1.82 mM in GM and WM across all subjects, respectively. Similarly, averaged ^2^H-water concentrations derived from FID-CRT DMI maps at ∼77 min after tracer administration were 15.18±3.16 mM and 12.46±1.57 mM in GM and WM, respectively. While no significant differences were detected between bSSFP-CRT and FID-CRT acquisition schemes in GM (*p*=0.73) and WM (*p*=0.96) regions, significantly higher ^2^H-water concentrations were observed in GM compared to WM dominated regions for both acquisition schemes (bSSFP-CRT: +27%, *p*=0.003; FID-CRT: +22%, *p*=0.004).

Regionally averaged ^2^H-Glc, ^2^H-Glx and ^2^H-water concentrations acquired in all volunteers at the last bSSFP-CRT time point (∼70min after tracer intake) and the last FID-CRT time point (∼77min after tracer intake) are shown in Table 2.

**Table 2:**
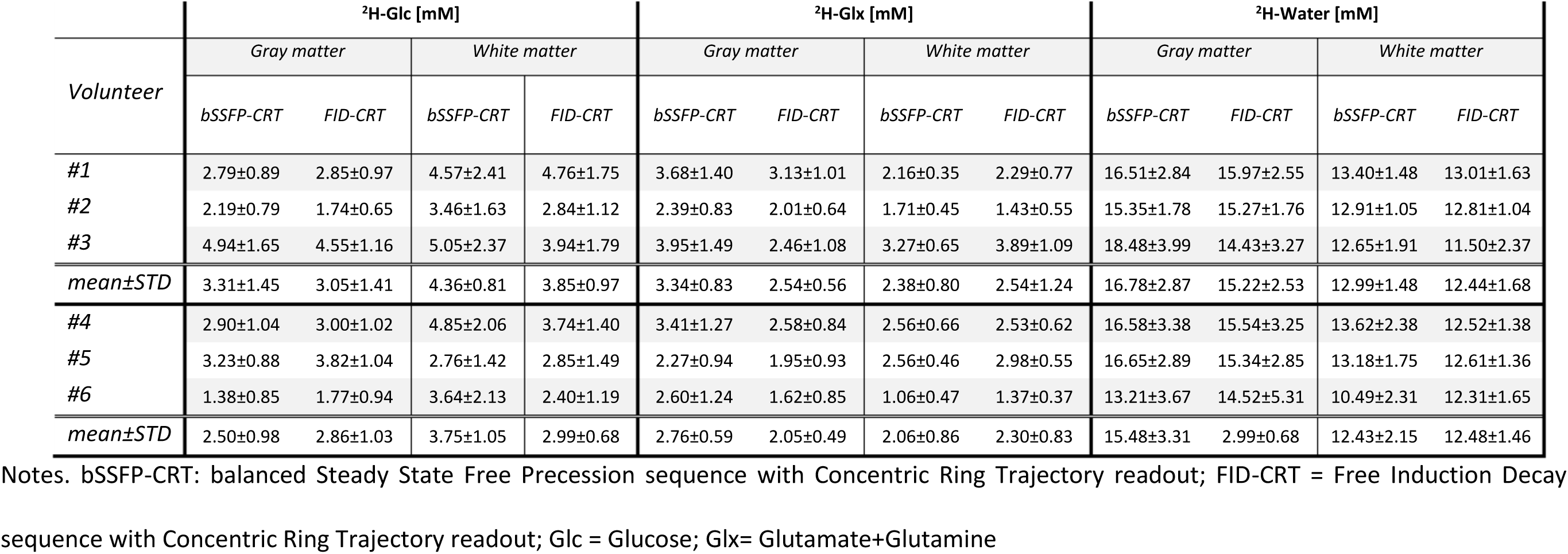
Individual concentrations for ^2^H-labeled glucose (Glc), glutamate+glutamine (Glx) and water from gray and white matter (GM/WM) dominated regions in the human brain, acquired 70 and 77min after oral tracer uptake using a balanced Steady State Free Precession sequence with Concentric Ring Trajectory readout (bSSFP-CRT) and a Free Induction Decay sequence with Concentric Ring Trajectory (FID-CRT) deuterium metabolic imaging (DMI). Subjects 1-3 were imaged at matching spatial resolution (0.75ml isotropic), subjects 4-6 with 0.36ml isotropic resolution in the bSSFP-CRT sequence and 0.75ml isotropic resolution in the FID-CRT sequence.

| Volunteer | <sup>2</sup>H-Glc [mM] |  |  |  | <sup>2</sup>H-Glx [mM] |  |  |  | <sup>2</sup>H-Water [mM] |  |  |  |
| --- | --- | --- | --- | --- | --- | --- | --- | --- | --- | --- | --- | --- |
|  | Gray matter |  | White matter |  | Gray matter |  | White matter |  | Gray matter |  | White matter |  |
|  | bSSFP-CRT | FID-CRT | bSSFP-CRT | FID-CRT | bSSFP-CRT | FID-CRT | bSSFP-CRT | FID-CRT | bSSFP-CRT | FID-CRT | bSSFP-CRT | FID-CRT |
| #1 | 2.79±0.89 | 2.85±0.97 | 4.57±2.41 | 4.76±1.75 | 3.68±1.40 | 3.13±1.01 | 2.16±0.35 | 2.29±0.77 | 16.51±2.84 | 15.97±2.55 | 13.40±1.48 | 13.01±1.63 |
| #2 | 2.19±0.79 | 1.74±0.65 | 3.46±1.63 | 2.84±1.12 | 2.39±0.83 | 2.01±0.64 | 1.71±0.45 | 1.43±0.55 | 15.35±1.78 | 15.27±1.76 | 12.91±1.05 | 12.81±1.04 |
| #3 | 4.94±1.65 | 4.55±1.16 | 5.05±2.37 | 3.94±1.79 | 3.95±1.49 | 2.46±1.08 | 3.27±0.65 | 3.89±1.09 | 18.48±3.99 | 14.43±3.27 | 12.65±1.91 | 11.50±2.37 |
| mean±STD | 3.31±1.45 | 3.05±1.41 | 4.36±0.81 | 3.85±0.97 | 3.34±0.83 | 2.54±0.56 | 2.38±0.80 | 2.54±1.24 | 16.78±2.87 | 15.22±2.53 | 12.99±1.48 | 12.44±1.68 |
| #4 | 2.90±1.04 | 3.00±1.02 | 4.85±2.06 | 3.74±1.40 | 3.41±1.27 | 2.58±0.84 | 2.56±0.66 | 2.53±0.62 | 16.58±3.38 | 15.54±3.25 | 13.62±2.38 | 12.52±1.38 |
| #5 | 3.23±0.88 | 3.82±1.04 | 2.76±1.42 | 2.85±1.49 | 2.27±0.94 | 1.95±0.93 | 2.56±0.46 | 2.98±0.55 | 16.65±2.89 | 15.34±2.85 | 13.18±1.75 | 12.61±1.36 |
| #6 | 1.38±0.85 | 1.77±0.94 | 3.64±2.13 | 2.40±1.19 | 2.60±1.24 | 1.62±0.85 | 1.06±0.47 | 1.37±0.37 | 13.21±3.67 | 14.52±5.31 | 10.49±2.31 | 12.31±1.65 |
| mean±STD | 2.50±0.98 | 2.86±1.03 | 3.75±1.05 | 2.99±0.68 | 2.76±0.59 | 2.05±0.49 | 2.06±0.86 | 2.30±0.83 | 15.48±3.31 | 2.99±0.68 | 12.43±2.15 | 12.48±1.46 |

Notes. bSSFP-CRT: balanced Steady State Free Precession sequence with Concentric Ring Trajectory readout; FID-CRT = Free Induction Decay
sequence with Concentric Ring Trajectory readout; Glc = Glucose; Glx= Glutamate+Glutamine

At matched spatial resolution both bSSFP-CRT and FID-CRT acquisition schemes yielded similar intra-subject coefficients of variation (CoV) for concentration estimates of ^2^H-Glc (GM: 34±2% vs 35±10%, WM: 21±5% vs 28±7%), ^2^H-Glx (GM: 49±3% vs. 43±7%, WM: 37±2% vs 40±10%) and ^2^H-water (GM: 19±5% vs. 21±9%, WM: 15±5% vs 13±4%). Furthermore, similar intra-subject CoV for concentration estimates of ^2^H-Glc (GM: 42±17%, WM: 29±14%) and ^2^H-Glx (GM: 51±8%, WM: 42±5%), as well as ^2^H-water Glc (GM: 22±5%, WM: 18±5%) were observed for bSSFP-CRT data with 2-fold increased spatial resolution.

### Comparison between DMI and CGM

After overnight fasting baseline glucose levels detected via the CGM sensor were 5.43±0.48 mM. 48±24 min after oral consumption of ^2^H-labeled glucose interstitial glucose level increased to 9.21±1.65 mM before returning to baseline levels. Individual time courses of interstitial glucose levels overlaid with brain ^2^H-Glc evolution detected via DMI are illustrated in Figure 5a-f.

**Figure 5:**
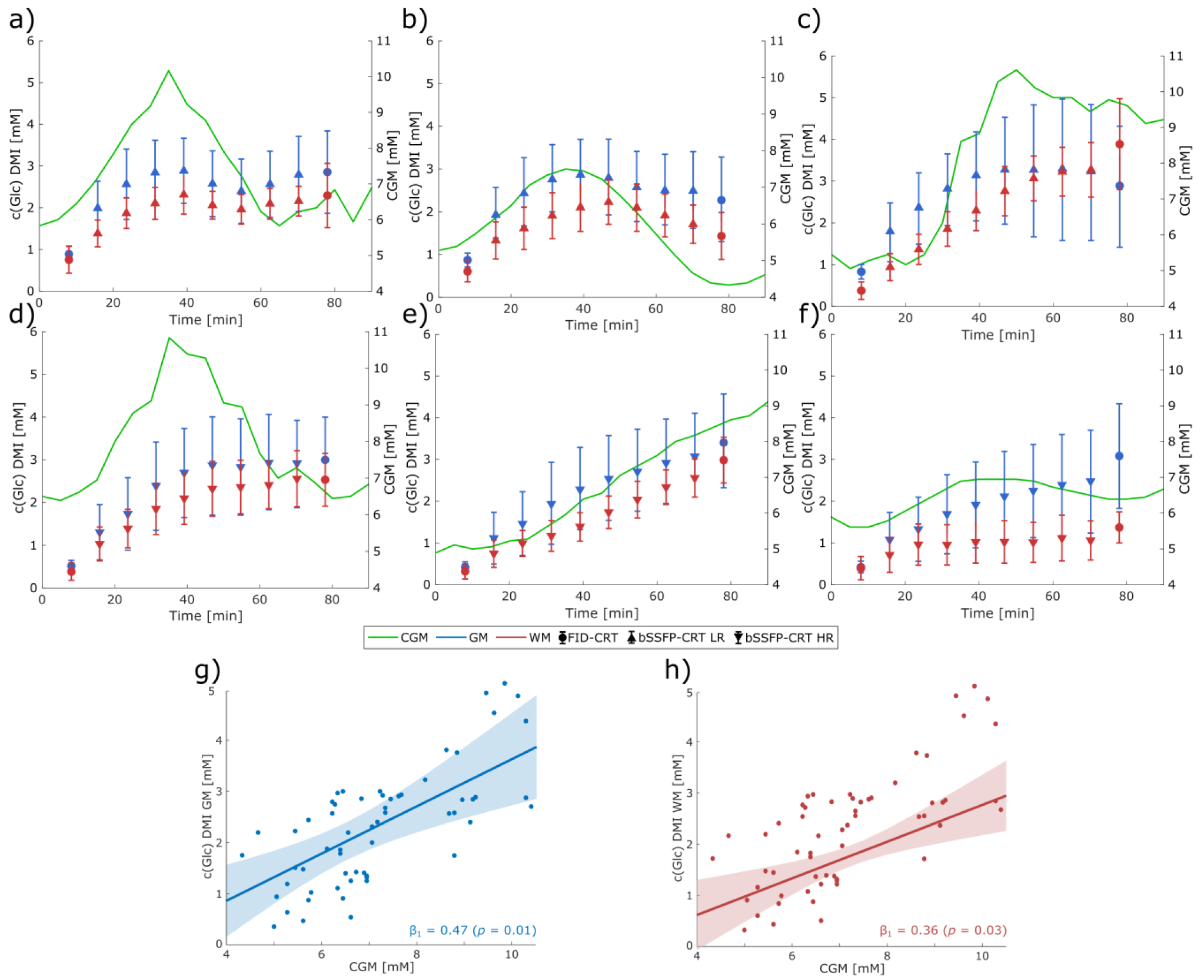
Glucose (Glc) levels measured in gray and white matter (GM in blue, WM in red) with Deuterium Metabolic Imaging (DMI; median and interquartile range) and in the interstitial fluid with a continuous glucose monitoring (CGM) sensor in mM in all six subjects (a-f). The relationships between GM/WM Glc levels and CGM Glc levels were analyzed using a linear mixed effects model, setting the CGM as fixed effect. Significant fixed effects were found for both GM (β_1_ = 0.47, *p* = 0.01) and WM (β_1_ = 0.36, *p* = 0.03) with no significant intercepts β_0_ (β_0,GM_ = −1.03, β_0,WM_ = −0.81).

The linear mixed models yielded a significant fixed effect of CGM in both GM (β_1_=0.47, *p*=0.01) and WM (β_1_=0.36, *p*=0.03) with no significant intercepts β_0_ (β_0,GM_ =-1.03 mM, β_0,WM_ =-0.81 mM), see Figure 5g-h. Inter-subject variability detected as intercept variance was found to be 2.66 mM in GM and 2.57 mM in WM.

The mean AUC of the interstitial fluid Glc measurements using a CGM sensor on the upper arm was 508.94±64.69 mM·min. In contrast, the mean AUC of dynamic brain Glc measurements using DMI was 148.53±43.38 mM·min, resulting in a mean ratio of 0.29±0.07 across all subjects.

## Discussion

This work presents a novel bSSFP acquisition scheme combined with a fast CRT readout for dynamic DMI measurements on human brains at 7T, with up to 3-fold increase in SNR compared to spoiled FID-MRSI acquisitions. This allowed us to improve the nominal spatial resolution ∼two-fold, compared to reported whole brain DMI maps acquired at comparable magnetic field strengths (13, 31).

^2^H-Glc uptake and downstream oxidative metabolism in the human brain could be observed dynamically in six healthy volunteers after oral administration of ^2^H-labeled Glc over ∼80 min by dynamically acquiring whole-brain metabolic maps of ^2^H-Glc, ^2^H-Glx and ^2^H-water with two different acquisition schemes, FID-CRT and novel bSSFP-CRT, at 0.75 ml and 0.36 ml isotropic resolution. Improved SNR of bSSFP-CRT acquisitions could potentially facilitate the observation of low metabolite concentrations, e.g., directly after tracer uptake in the beginning of the experiment, and in smaller brain structures. This is more challenging using less sensitive spoiled FID-CRT acquisitions, where quantification in many voxels fails and need to be excluded as reported in a previous study (13). In addition, the metabolic maps acquired with 2-fold higher spatial resolution using the bSSFP-CRT sequence did not negatively impact the concentration estimation of ^2^H-Glc and ^2^H-Glx and featured similar results with comparable regional CoV compared to acquisitions with lower isotropic resolution. Furthermore, the signal distribution is more homogeneous in GM and WM tissue for 3D DMI maps of ^2^H-Glc and ^2^H-Glx and ^2^H-water acquired with bSSFP-CRT than for FID-CRT, allowing better delineation of the tissue types.

Despite different T_1_-weighting effects between the two acquisition schemes, similar concentration estimates were obtained in GM and WM for all three metabolites, ^2^H-Glc, ^2^H-Glx and ^2^H-water approximately 70 min after tracer uptake, and are in line with previous studies (9, 11, 13, 35). Lower ^2^H-Glx concentrations in WM dominated regions were derived from FID-CRT data leading to a higher GM/WM contrast compared to bSSFP-CRT acquisitions, while similar GM/WM contrast was observed for ^2^H-Glc concentration. This could be presumably explained by prolonged T_1_ relaxation times of ^2^H-Glx (T_1_=165 ms) versus ^2^H-Glc (T_1_=70ms) (9, 12, 31), and by the differences in contrast observed in the metabolic maps. In those, an increased ^2^H-water contrast was observed with the bSSFP-CRT sequence compared to the ^2^H-water maps acquired using FID-CRT, while ^2^H-Glc contrast was very similar and contrast in ^2^H-Glx varied between subjects.

Dynamic Glc measurements of the brain via DMI were complemented with simultaneous minimally invasive continuous monitoring of systemic Glc levels of the interstitial tissue on the upper arm using an MR-safe CGM sensor (20), which is commonly used in diabetes care. As inter-subject variability is relatively high for metabolic studies using oral glucose administration, this simple and affordable approach could be helpful as an additional reference for normalization. Interstitial Glc values are not equivalent to assessments of venous or capillary blood glucose levels, as has been done previously (36–39), but would be desirable to fully understand the underlying metabolic model of brain glucose metabolism.

This would require consecutive blood sampling and estimation of deuterium enrichment to derive real quantitative turnover rates and metabolic fluxes, which was not the aim of this study. Consequently, we did not include invasive blood sampling in our study protocol.

Despite the small sample size, we were able to show a significant association between brain Glc uptake and systemic Glc levels as measured simultaneously in interstitial fluid with a CGM sensor. As seen in Figure 5a-f, the dynamics of ^2^H-Glc measured in the brain using DMI follow the dynamics of the systemic Glc measured with CGM in most subjects, but at lower levels and with blunted peaks. Similar associations between brain and plasma Glc have been shown in single voxel ^1^H-MRS studies (40, 41) and is in excellent agreement with brain Glc uptake being driven mainly by a concentration gradient (1).

Interestingly, while the AUCs from the CGM curve and the DMI curves have very different physical origins and vary among individuals, the 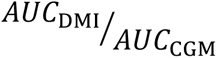 ratio was very similar when considering all participants. Interpretation of this ratio, however, would require a larger and more diverse sample combined with better metabolic characterization of the participants. Complementing DMI with CGM measurements also allowed us to observe systemic Glc changes over a period of up to 14 days. This could enable a more thorough investigation of the interplay between systemic metabolic aberrations and changes in brain metabolism.

The 3-fold SNR gains of ^2^H-water and ^2^H-Glc resonances of the novel bSSFP-CRT sequence compared to spoiled FID-MRSI acquisitions that we obtained in phantom experiments, are in line with a previous preclinical study (17). Our initial phantom was made of three cylinders, which contained natural abundance levels of ^2^H-Glc, ^2^H-water and ^2^H-Lac. However, the oily lactate solution used in our study proved to be challenging as strong B_0_ inhomogeneities could not be sufficiently corrected. Consequently, we were not able to reliably estimate ^2^H-Lac signals using both acquisitions schemes to determine the SNR gain.

Although parameter estimation for bSSFP acquisitions included optimization for ^2^H-Lac signals, we were not able to reliably detect increasing ^2^H-Lac levels *in vivo*. An explanation could be that only ∼15% of glucose is anaerobically converted to lactate in the healthy human brain (1), which is still below the detection limit of our method. This means that further optimizations are needed to obtain reliable dynamic ^2^H-Lac maps using bSSFP-CRT. These might in turn enable the estimation of glycolytic flux rates in pathologies with increased lactate synthesis, e.g., tumors with increased Lac levels are due to the Warburg effect (3, 9, 42). Including such measurements of participants with brain tumors was outside the scope of this feasibility study but is to be performed in future studies. Furthermore, we could not determine the SNR gain for ^2^H-Glx during phantom measurements, as the cost of this deuterated tracer was unjustifiably high.

While *in vivo* determination of SNR differences between FID-CRT and bSSFP-CRT sequences is theoretically possible, unstable metabolic concentrations and the long acquisition times of these repetitive scans, complicated these measurements. For further protocol optimization, a more realistic brain phantom should be used.

Although the developments here introduced allowed us to increase spatial resolution by 2-fold, this is still comparably low to established metabolic imaging modalities such as FDG-PET. This is still a major limitation of DMI and hinders translation into clinical application. Further SNR increases via postprocessing steps such as denoising algorithms (13, 14, 18, 43) might facilitate higher spatial resolution in future studies when combined with bSSFP-CRT techniques. This would improve the point spread function and significantly reduce partial volume errors, possibly allowing the observation of more local metabolic differences, and thus, paving the way for clinical application of DMI.

## Conclusion

This study demonstrates the feasibility of dynamic deuterium metabolic imaging to non-invasively map whole-brain glucose metabolism with up to 3-fold increased signal-to-noise ratio and 2-fold increase of spatial resolution using a novel balanced Steady State Free Precession (bSSFP) pulse sequence combined with fast spatial-spectral k-space sampling at 7T. Dynamic brain glucose concentrations were complemented by simultaneously measuring systemic glucose levels in interstitial fluid minimally invasively using a continuous glucose monitoring sensor that is used in diabetes care.

This suggests significant potential for clinical applications with improved characterization of local pathologic brain metabolism in conjunction with systemic metabolic aberrations in a minimally invasive way.

## Acknowledgements

The financial support by the Austrian Federal Ministry for Digital and Economic Affairs, the National Foundation for Research, Technology and Development and the Christian Doppler Research Association, Austrian Science Fund and National Institute of Health is gratefully acknowledged. This research was funded in whole or in part by the Austrian Science Fund (FWF) [10.55776/KLI1106]. For open access purposes, the author has applied a CC BY public copyright license to any author accepted manuscript version arising from this submission. Funded by the European Union (ERC, GLUCO-SCAN, 101088351). Views and opinions expressed are however those of the author(s) only and do not necessarily reflect those of the European Union or the European Research Council Executive Agency. Neither the European Union nor the granting authority can be held responsible for them.

## Funding

This work was supported by the National Institute of Health NIH R01EB031787, the Austrian Science Fund: WEAVE I 6037 & KLI 1106, the Christian Doppler Laboratory for MR Imaging Biomarkers (BIOMAK) and the European Union (ERC, GLUCO-SCAN, 101088351). The Weizmann coworkers were supported by the Israel Ministry of Health, and by the Israel Cancer Research Foundation.

## Conflicts of interest

R. Lanzenberger received investigator-initiated research funding from Siemens Healthcare regarding clinical research using PET/MR. He is a shareholder of the start-up company BM Health GmbH since 2019.

## Abbreviations

AUC: Area Under the Curve
BMI: Body Mass Index
bSSFP: balanced Steady State Free Precession
bSSFP-CRT: balanced Steady State Free Precession combined with concentric ring trajectories
CGM: Continuous Glucose Monitoring
CoV: Coefficient of Variation
CRLB: Cramer Rao Lower Bound
CRT: Concentic Ring Trajectory
CSF: Cerebrospinal Fluid
DMI: Deuterium Metabolic Imaging
ΔT_E_: Echo spacing
FDG-PET: [^18^F]-Fluorodeoxy Glucose Positron Emission Tomography
FID: Free Induction Decay
FID-CRT: Free Induction Decay Concentric Ring Trajectory
Glc: Glucose
Glx: Glutamate+Glutamine
GM: Gray Matter
IDEAL: Iterative Decomposition of water and fat with Echo Asymmetric and Least-squares estimation
Lac: Lactate
MRI: Magnetic Resonance Imaging
MRS/MRSI: Magnetic Resonance Spectroscopy/Spectroscopic Imaging
SNR: Signal-to-noise ratio
STD: Standard deviation
TCA: Tricarboxylic acid cycle
T_R_: Repetition time
WM: White Matter

## Supplemental Digital Content

**Supplemental Digital Content Table 1:** Minimum Reporting Standards for in vivo MR Spectroscopy Note. – Parameters 7T DMI, CRLB = Cramér-Rao lower bounds; FID = free induction decay; CRT = concentric ring trajectory; FOV = field of view; FWHM = full-width-at-half-maximum; Glx = Glutamate+Glutamine; Glc = Glucose; SNR = signal-to-noise ratio; VOI = volume of interest.

| Minimum Reporting Standards in MR Spectroscopy (according to Lin et al. NMR Biomed 2021) |  |
| --- | --- |
| **1. Hardware** |  |
| a. Field strength [T] | 7T |
| b. Manufacturer | Siemens |
| c. Model (software version if available) | Magnetom dot Plus |
| d. RF coils: nuclei (transmit/ receive), number of channels, type, body part | <sup>2</sup>H/<sup>1</sup>H dual tuned quadrature birdcage head coil, transmit/receive, 1 channel, (Stark Contrast MRI Coils Research, Germany) |
| e. Additional hardware | N/A |
| **2. Acquisition** |  |
| a. Pulse sequence | 3D FID-acquire density-weighted concentric ring trajectory (CRT) MRSI & 3D multi-echo balanced Steady State Free Precession (bSSFP) density-weighted concentric ring trajectory (CRT) MRI |
| b. Volume of Interest (VOI) locations | whole-brain, unlocalized excitation using rectangular RF pulse with 86° (FID-CRT) & ±50° (bSSFP-CRT) flip angle |
| c. Nominal VOI size [cm<sup>3</sup>, mm<sup>3</sup>] | FOV 200x200x192 mm<sup>3</sup> |
| d. Repetition Time (TR), Echo Time (TE) [ms, s] | FID-CRT: TR=290 ms / 2 ms acquisition delay<br>bSSFP-CRT: TR = 23 ms / 2 ms acquisition delay |
| e. Total number of Excitations or acquisitions per spectrum | FID-CRT: 43 circles (7 min acquisition time)<br>bSSFP-CRT: 107 circles, 2 averages (matched 7 min acquisition time) |
| In time series for kinetic studies | N/A |
| i. Number of Averaged spectra (NA) per time-point | N/A |
| ii. Averaging method (e.g. block-wise or moving average) | N/A |
| iii. Total number of spectra (acquired / in time-series) | N/A |
| f. Additional sequence parameters (spectral width in Hz, number of spectral points, frequency offsets); If STEAM: Mixing Time TM; If MRSI: 2D or 3D, FOV in all directions, matrix size, acceleration factors | FID-CRT: BW: 380 Hz, 96 spectral points, 22x22x21<br>bSSFP-CRT: BW: 285 Hz, 5 echoes, 22x22x21 & 28x28x27 |
| g. Water Suppression Method | No water suppression |
| h. Shimming Method, reference peak, and thresholds for “acceptance of shim” chosen | Standard shim + manual adjustment, <sup>1</sup>H water peak < 40 Hz, <sup>2</sup>H water peak < 30 Hz Region: whole brain |
| i. Triggering or motion correction method (respiratory, peripheral, cardiac triggering, incl. device used and delays) | - |
| **3. Data analysis methods and outputs** |  |
| a. Analysis software | FID-CRT: LCModel 6.3-1<br>bSSFP-CRT: inhouse developed k-space IDEAL algorithm in MATLAB 2017 |
| b. Processing steps deviating from quoted reference or product | N/A |
| c. Output measure (e.g. absolute concentration, institutional units, ratio) | concentration estimation in mM |
| d. Quantification references and assumptions, fitting model assumptions | Simulated in NMRScope-B |
| **4. Data Quality** |  |
| a. Reported variables (SNR, Linewidth (with reference peaks)) | SNR was calculated using voxel-wise mean and standard deviation across 5 repeated measurements |
| b. Data exclusion criteria | CRLBs > 50 % for water, Glc, Glx, no CRLB threshold for first time point |
| c. Quality measures of postprocessing Model fitting (e.g. CRLB, goodness of fit, SD of residual) | CRLB for FID-CRT datasets<br>None for bSSFP-CRT datasets |
| d. Sample Spectrum | - |

**Supplemental Digital Content Figure 1:**
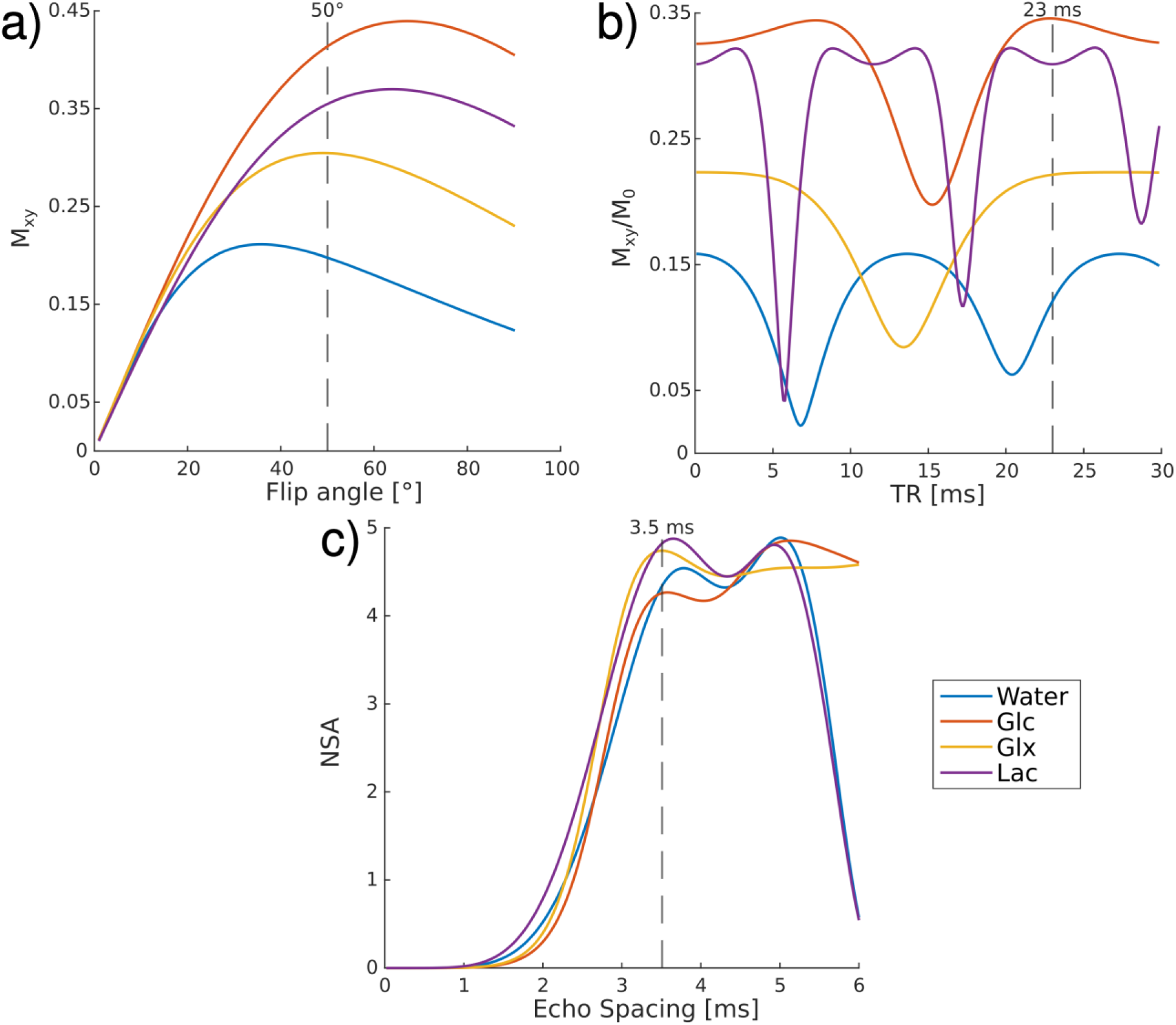
Parameter optimization according to D. Peters et al., 2021, and pulse design of the balanced Steady State Free Precession sequence combined with concentric ring trajectory readout.

a. Calculation of optimal flip angle for the metabolites of interest, ^2^H-Glucose (Glc), ^2^H-Glutamate+Glutamine (Glx), ^2^H-Lactate (Lac) and ^2^H-Water. As a compromise between signal gain of all metabolites and limitation of Specific Absorption Rate (SAR), a flip angle of 50° was chosen.
b. Repetition time (T_R_), calculated with A2, was set to 23ms to avoid banding artifacts for all metabolites, and accommodate 5 echoes for metabolite separation using an IDEAL algorithm.
c. Echo spacing (ΔT_E_) was set to 3.5ms to optimize phase difference between the four metabolites of interest for optimal metabolite separation using an IDEAL algorithm, expressed with the number of signal averages(NSA), while keeping T_R_ as short as possible for 5 echoes.

**Supplemental Digital Content Figure 2:**
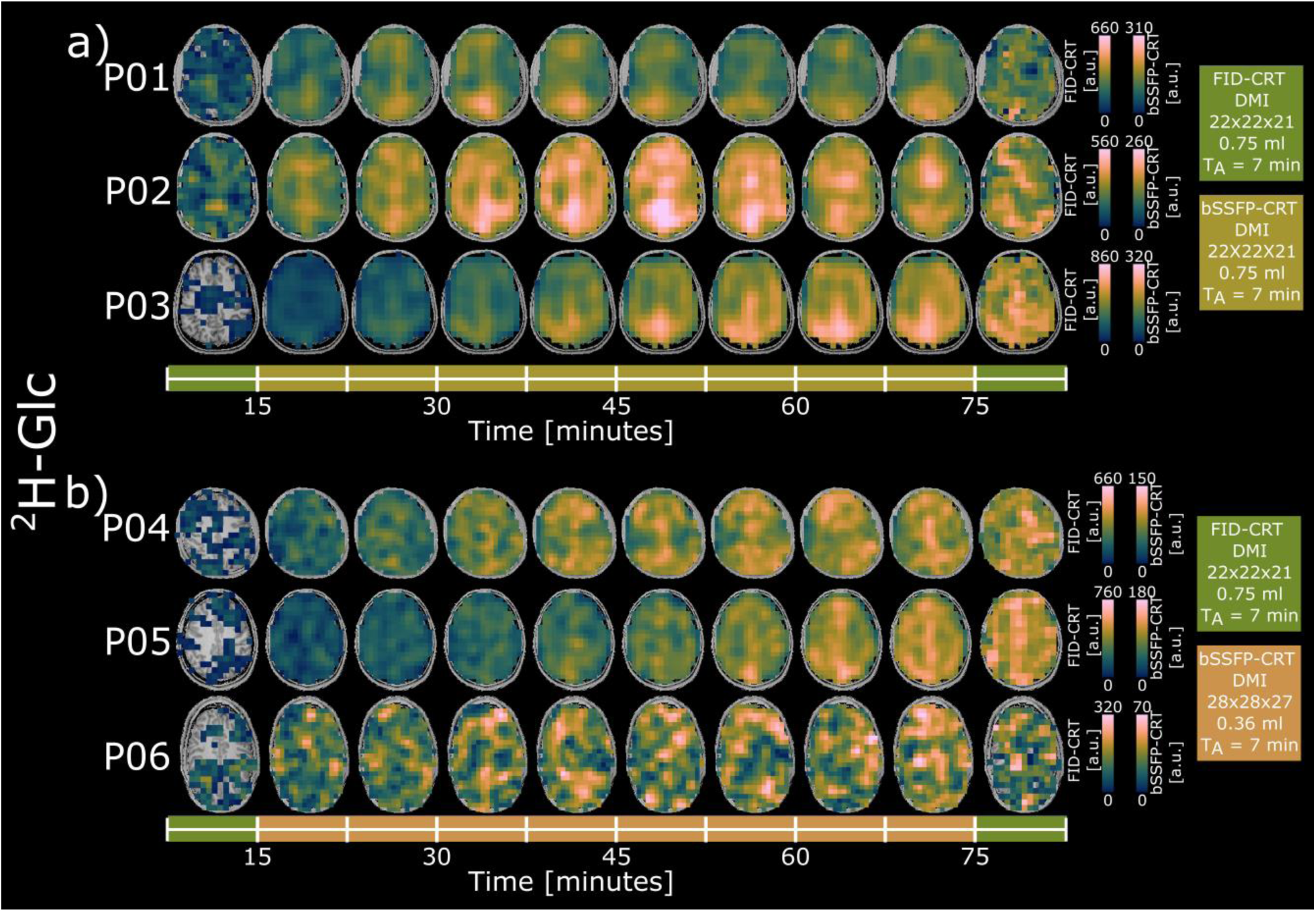
Time courses of axial ^2^H-Glucose (Glc) maps given in arbitrary units (a.u.) from all participants (a: both acquisitions 0.75ml isotropic resolution, b: FID-CRT 0.75ml, bSSFP-CRT 0.36ml isotropic resolution), detected using deuterium metabolic imaging (DMI) with the Free Induction Decay Concentric Ring Trajectory (FID-CRT, green) and the balanced Steady State Free Precession Concentric Ring Trajectory (bSSFP-CRT, olive green and orange) sequences at 7T. Missing voxels in the metabolic maps do not contain a value (NaN: not a number).

**Supplemental Digital Content Figure 3:**
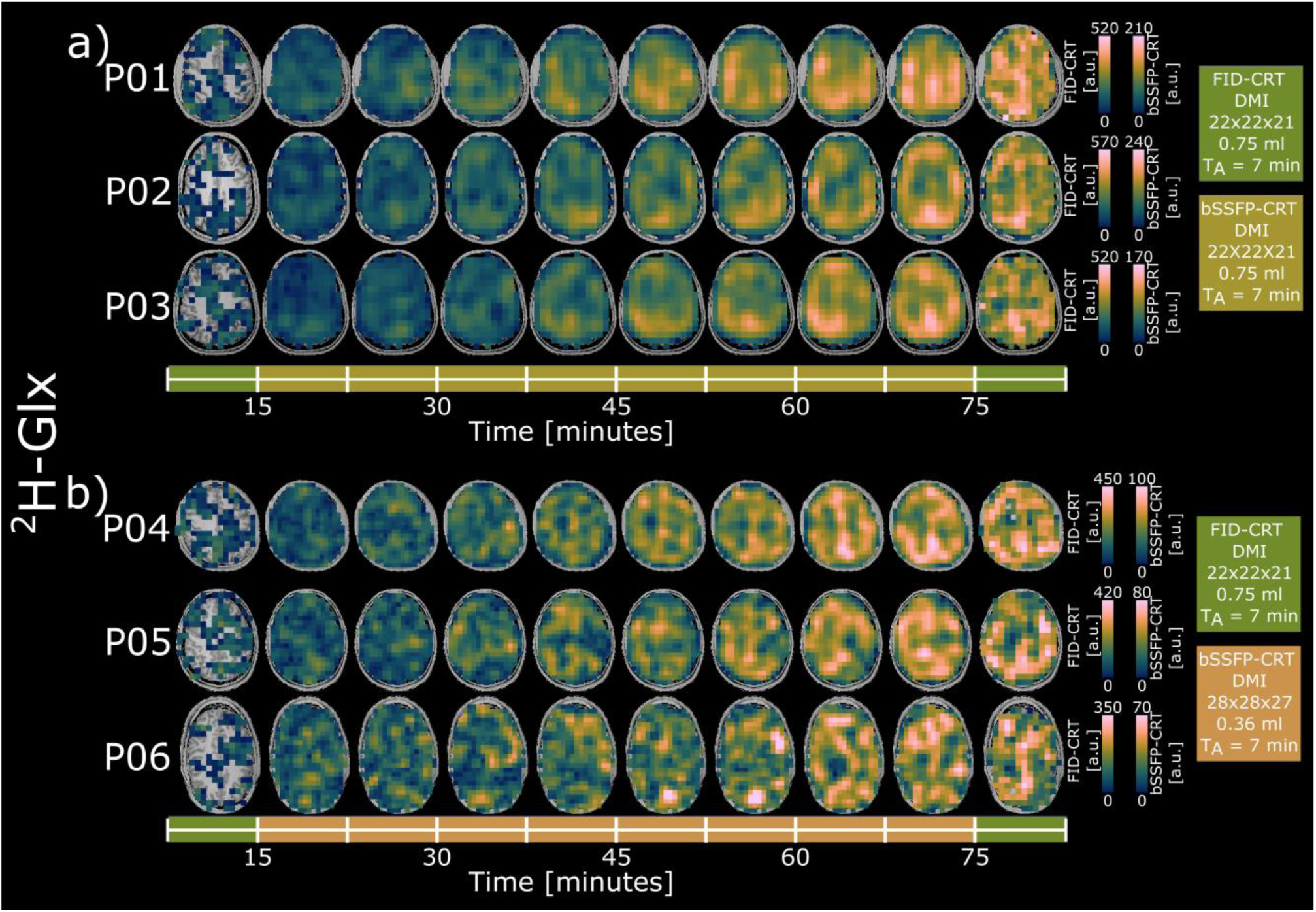
Time courses of axial ^2^H-labeld Glutamate+Glutamine (Glx) maps given in arbitrary units (a.u.) from all participants (a: both acuqisitions 0.75ml isotropic resolution, b: FID-CRT 0.75ml, bSSFP-CRT 0.36ml isotropic resolution), detected using deuterium metabolic imaging (DMI) with the Free Induction Decay Concentric Ring Trajectory (FID-CRT, green) and the balanced Steady State Free Precession Concentric Ring Trajectory (bSSFP-CRT, olive green and orange) sequences at 7T. Missing voxels in the metabolic maps do not contain a value (NaN: not a number).

**Supplemental Digital Content Figure 4:**
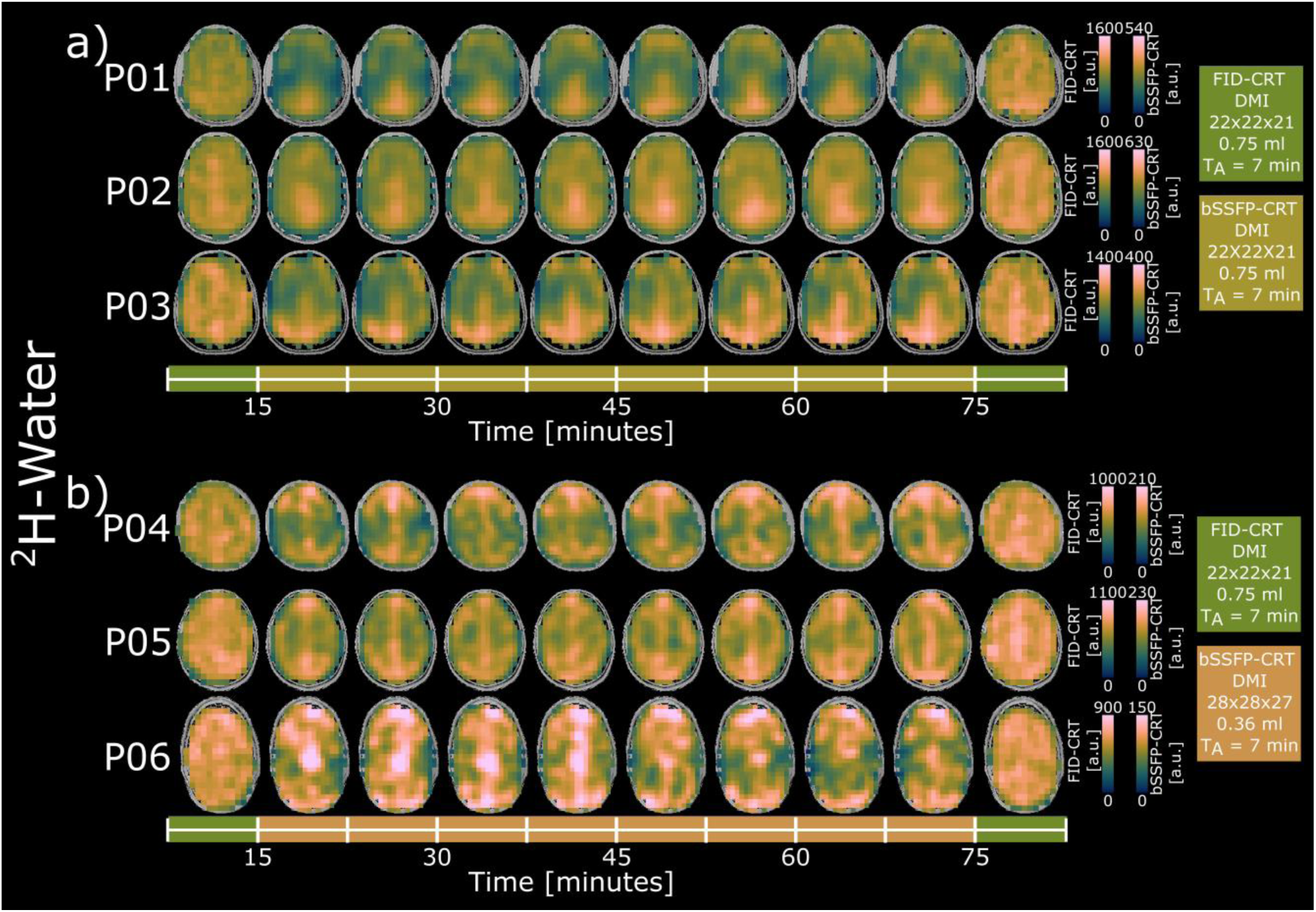
Time courses of axial ^2^H-water maps given in arbitrary units (a.u.) from all participants (a: both acquisitions 0.75ml isotropic resolution, b: FID-CRT 0.75ml, bSSFP-CRT 0.36ml isotropic resolution), detected using deuterium metabolic imaging (DMI) with the Free Induction Decay Concentric Ring Trajectory (FID-CRT, green) and the balanced Steady State Free Precession Concentric Ring Trajectory (bSSFP-CRT, brown and orange) sequences at 7T.

## Appendix 1

Absolute concentration estimation in mM units of ^2^H resonances detected using Deuterium Metabolic Imaging (DMI) at 7T with Free Induction Decay Concentration Ring Trajectory (FID-CRT) and balanced Steady State Free Precession Concentric Ring Trajectory (bSSFP-CRT). The amplitude of the deuterated metabolites was referenced voxel wise to the amplitude of the deuterated water signals. The amplitude ratios were corrected for the relaxation factors (equations A1 and A2) and fractional water content for gray and white matter (GM, WM) and cerebrospinal fluid (CSF) and tissue water content with *d*_*GM*_=0.78, *d*_*WM*_=0.65 and *d*_*CSF*_=0.97. Fractional water content of CSF was excluded for ^2^H-Glx concentration estimation. Label loss of ^2^H-Glx was corrected according to de Graaf et al., ACS Chemical Neuroscience, 2021 (assuming on average approximately 40%).

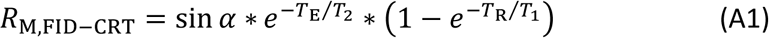

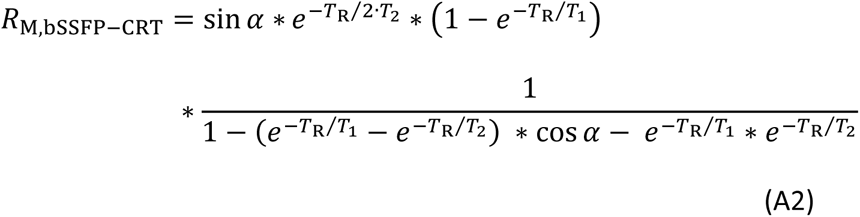

## Notes

### Author Declarations

The study was approved by the local ethics committee of the Medical University of Vienna according to the guidelines of the Declaration of Helsinki.

